# Novel Electrode Architecture for Subgaleal Electroencephalography: A Feasibility Study

**DOI:** 10.1101/2021.08.25.21262594

**Authors:** Arman Ahnood, Nhan Duy Truong, Bobbi Fleiss, Armin Nikpour, Omid Kavehei

## Abstract

Electroencephalography (EEG) has been widely used to understand the nervous system and as a clinical diagnostic tool. In the case of neurological conditions with intermittent episodes, such as epilepsy, long-term EEG monitoring outside the clinics and in the community setting is vital. Subgaleal EEG (sgEEG) has emerged as an essential tool for long-term monitoring over several years. Current sgEEG solutions share a need for at least a 10 cm long lead wire, resulting in a bulky and invasive device. This work introduces a novel electrode architecture for subgaleal EEG recording, which forgoes the need for lead wires. A back-to-back electrode configuration with an electrode spacing of less than 1 mm is proposed. Compared to the current side-by-side approaches with an electrode spacing of several cm, our proposed approach results in at least one order of magnitude reduction in volume. The efficacy of the proposed electrode architecture is investigated through finite element modeling, phantom measurements, and cadaver studies. Our results suggest that compared to the conventional side-by-side electrode configuration, the source signal can be recorded reliably. Lead wires have posed a significant challenge from a device reliability and measurement quality perspective. Moreover, lead wires and the associated feedthrough connectors are bulky. Our proposed lead-free EEG recording solution may lead to a less invasive surgical placement through volume reduction and improve EEG recording quality.

## 1 Introduction

Epilepsy is one of the most common, complex, and widely misunderstood neurological disorders, impacting people of all ages and genders [1]. Epilepsy causes unprovoked seizures, which can briefly disturb the brain’s electrical activity and cause temporary interruption or changes in bodily functions, movement, awareness, behavior, or feelings [1]. Objective counting of seizures is essential in the diagnosis and treatment of epilepsy [2]. Epilepsy affects about one percent of the world’s population [3]. About 30% to 35% of people living with epilepsy are diagnosed with uncontrolled or partially controlled seizures. The road to this diagnosis is long and cumbersome. The data shows the efficacy of anti-epileptic drugs (AEDs) in achieving seizure freedom, reduces from around 45% with the trial of the first AED to just 7% for the trial of the third AED [1, 4, 5]. Only a proportion of patients that are resistant to AED, may be candidates for surgery or neuromodulation such as vagus nerve stimulation (VNS). These patients are left with no other therapeutic options to achieve seizure control [3, 4]. Poor seizure control is more prevalent in people living with epilepsy who live in low- and middle-income countries where there is no or limited access to proper medical care [6]. While in most epilepsy patients, a cause cannot be identified, known aetiologies include genetic, trauma, brain tumors, stroke, Alzheimer’s disease, and infection.

The golden standard for the diagnosis of epilepsy is surface electroencephalography (EEG) [8]. Since the 1920s, EEG has provided a tool to study and understand the neurophysiology of the human brain. EEG use in clinical practices, research institutions, at-home ambulatory service, and telehealth is snowballing [9]. Modern electronics, advancements in microelectronics, and the artificial intelligence revolution provide an accelerated drive to push for non-conventional, reliable, and long-term use of EEG, in particular for early seizure detection and prediction in uncontrolled epilepsy [10, 11, 12]. There has been a growing interest in long-term home-based ambulatory monitoring of epilepsy using surface EEG [13]. One reason has been the lack of objective data for seizure occurrence, which is used to diagnose and manage patients. Continuous monitoring of individuals as an inpatient is resource-intensive and costly.

Many patients note their seizures on paper diaries. These records have been shown to be inaccurate in studies. Part of the reason may be that the memory of patients during and after a seizure is impaired [14, 15]. Recordings using surface electrodes, in both dry or wet configurations, are considered the least invasive method. However, these approaches are accompanied by a poor signal to noise due to the unstable electrode-tissue interface. While the surface electrodes are well suited for short EEG recording sessions, they are not suitable for long-term EEG monitoring. Several alternative solutions have been proposed to address this problem. Among these are intracranially implanted electrodes, such as the ones used for electrocorticography (ECoG) [16]. This approach yields superior recording quality, with a very high signal-to-noise ratio (SNR) and long-term electrode stability. However, intracranially implanted electrodes are invasive and have a significant risk of complications [17, 16].

In the recent years, subgaleal EEG (sgEEG) has emerged as a possible candidate for long-term EEG monitoring [7, 18, 19, 20, 21, 22, 23, 24, 25, 26, 27, 28]. High correlation between scalp and subgaleal EEG recordings have also been reported [25, 29]. It has also been reported that a single extracranial electrode could maintain coherence with an intracranial EEG over a cortical field of view as significant as 150 cm^2^ [30]. It is also known that the very high ratio of dural potentials over scalp potential drops rapidly as a function of the area of the dipole layer on an idealized smooth model of the cortex [31]. A minimum of 6 cm^2^ synchronous cortical activity is necessary for epileptic rhythms to be recorded on the scalp [31].

There is also emerging evidence to support the need for chronic EEG monitoring to improve the care of patients with epilepsy [32]. Like intracranial electrodes, the implanted subgaleal electrodes provide a stable electrode-tissue interface, which is an essential requirement for reliable long-term EEG monitoring. Moreover, the placement of subgaleal electrodes under the scalp is far less invasive than an intracranial placement. This will lead to wider clinical adoption and clinical utility [7]. The current generation of EEG recording systems that utilize subscalp electrodes typically consist of two or more electrodes connected to the main body of the implant using lead wires (see Fig. 1(a)). These recording electrodes are positioned between the scalp and the skull. The implant body hosts the electronics in a hermetically sealed package along with wireless power/data telemetry. Electronic housing is re-purposed from implantable hearing aids in most current solutions and is relatively thick for a subgaleal or subscalp placement. These systems operate in the differential mode, meaning that the difference in the voltage between the two recording electrodes is measured. This requires the recording electrodes to be positioned such that the voltage sensed at the two electrodes is different. The current generation of devices achieves this by placing the electrodes at various locations from each other. The tissue impedance attenuates the electrical signal generated by the brain and sensed by the channels. This results in a difference in the voltage sensed by the two electrodes.

**Figure 1:**
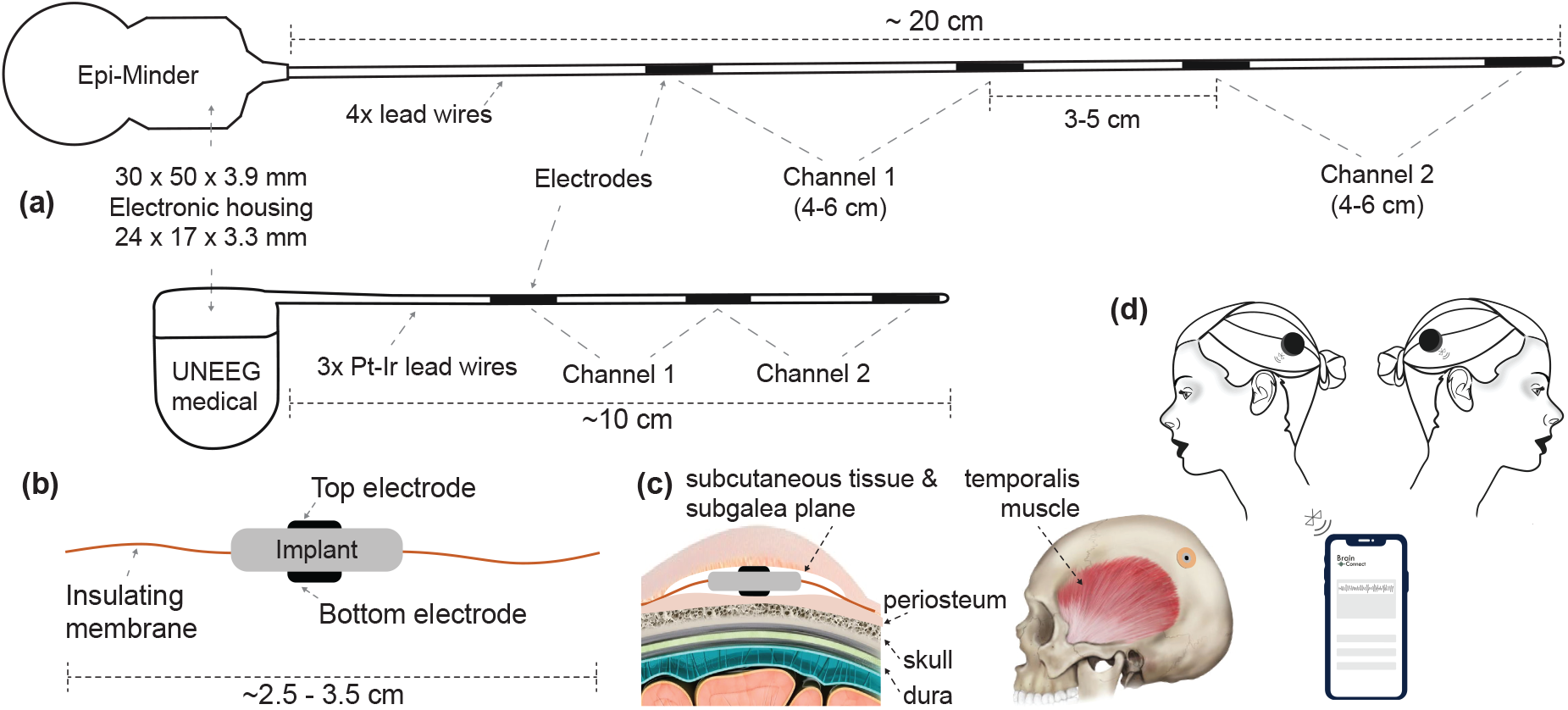
Current solutions for subscalp and subcutaneous EEG and our proposed solution. (a) Illustrates two examples of subscalp and subcutaneous technologies. Epi-Minder bilateral subscalp EEG device with four lead wires, with the longest being more than 20 cm. The Epi-Minder’s implantable body is comprised of four electrodes positioned along a length of the implantable body and using a cochlear implant system housing. Note that the channel arrangements may be different from one that is shown here. The leading solution from UNEEG medical’s unilateral SubQ device with two channels, a hearing-aid electronic housing, and three electrodes. Here we only show the implantable parts [7]. We estimate the Minder’s and SubQ’s implant volumes at *∼*6,000 *µ*l (with *∼*96% rigid parts) and *∼*1,500 *µ*l (with *∼*92% rigid parts), respectively. (b) Shows a simplified illustration of our proposed solution with novel electrode architecture. The two electrodes are placed on the top and bottom of the implant body, where electronic housing is. The insulating membrane, such as medical-grade silicone, extends around the rigid core of the implant’s body. The insulating membrane helps to achieve a few cm *sensing distance* with sub-mm *physical distance* between the two electrodes. We estimate the total implant volume to be *∼*1,000 *µ*l (with *∼*32% rigid parts), with an outer insulating membrane to be flexible and foldable when necessary. (c) Shows how the implant body subgaleal and possible positioning of the device on a human head. (d) Illustrates the external units. One of each side and uniquely tagged for communication.

One challenge of current subgaleal recording systems is a need for *>*10 cm long lead-wires. In the most recent design, the Epi-Minder device, the length of lead wires approaches 20 cm, and there is a need for four leads to establish two sgEEG recording channels that are implanted under general anesthesia [7] (see Fig. 1(a)). Lead wires are fragile conductive elements, and some implants must have special material and structure considerations such as a spiral shape to reduce the likelihood of breakage as the implant body stretches, bends, or flexes. They are manufactured using special materials such as platinum-iridium (Pt-Ir) alloy or titanium alloy. Laser welding is used to electrically and mechanically couple each electrode to its lead wire. Beyond the challenges associated with manufacturing the leads and their placement, the presence of lead wires introduces electrical interference noise and motion artifacts [20].

Even though implanted wires have been used since the 1960’s and supported by a multi-billion-dollar industry, the challenges outlined above remain. There are many examples of leadless implants that are more reliable and acceptable in the industry [33]. One main reason for a push for lead-less implants is repeated recalls, such as the recent FDA Class I recall of 20,000 implantable cardioverter-defibrillators (Boston Scientific - EMBLEM S-ICD) due to lead wire fracture in 2020 [34]. Lead-less implantable technology would be a significant improvement.

It is reported that typical complications of medical implants such as hematomas and fibrosis are rare in subgaleal electrodes [7]. The safety of subgaleal electrode placement has been reported, as has the efficacy of subscalp and subgaleal electrophysiological signal recordings [35, 18, 7]. Despite the skull attenuating high-frequency EEG signals, it has been reported that these electrodes can also record high-frequency signals (high-gamma) without frequency distortion [35, 36]. These high-frequency oscillations (HFOs) are becoming increasingly important in our understanding of seizures, seizure detection, and localization. While the spatial coverage in subgaleal EEG is currently significantly lesser than surface EEG, their higher reliability is shown to result in improved signal quality [18]. This opens up an exciting opportunity for a widespread and reliable diagnostic tool that has a primary focus on epilepsy.

We present the feasibility study on whether the electrode placement on the top and bottom plates of an implant body’s top can obtain EEG signals reliably. An insulating membrane is used to enhance the differential recording between two back-to-back electrodes (see Fig. 1(b, c, and d)). Using electrostatic finite-element modeling as well as experiments using phantom and animal cadaver models, we investigate the capabilities and limitations of the proposed approach. As well as the benefits of not having lead wires, this approach enables a substantial reduction in the implant size, allowing minimally invasive placement under local anesthesia. With this design, the electrodes are sub-mm *physical distance* but a few cm *sensing distance* from each other. This is due to a non-conductive and elastomeric material extension around the rigid core, such as medical-grade silicone. This is the first time in this context where the *physical* and *sensing* distances are vastly different to the best of our knowledge. Our modeling and experimental results have shown that we should be able to record EEG reliably with an implant of a few centimeters in diameter. In this feasibility study, we present modeling and experimental evidence that the back-to-back electrode architecture is feasible.

## 2 Methods

### 2.1 Electrostatic finite element modelling

A volume conduction model consisting of a seven-layered sphere was used to represent a human head. The electrical properties of the layers, as well as their sizes, were adopted from [37] and are summarised in Table 2.1.

**Table 1:**
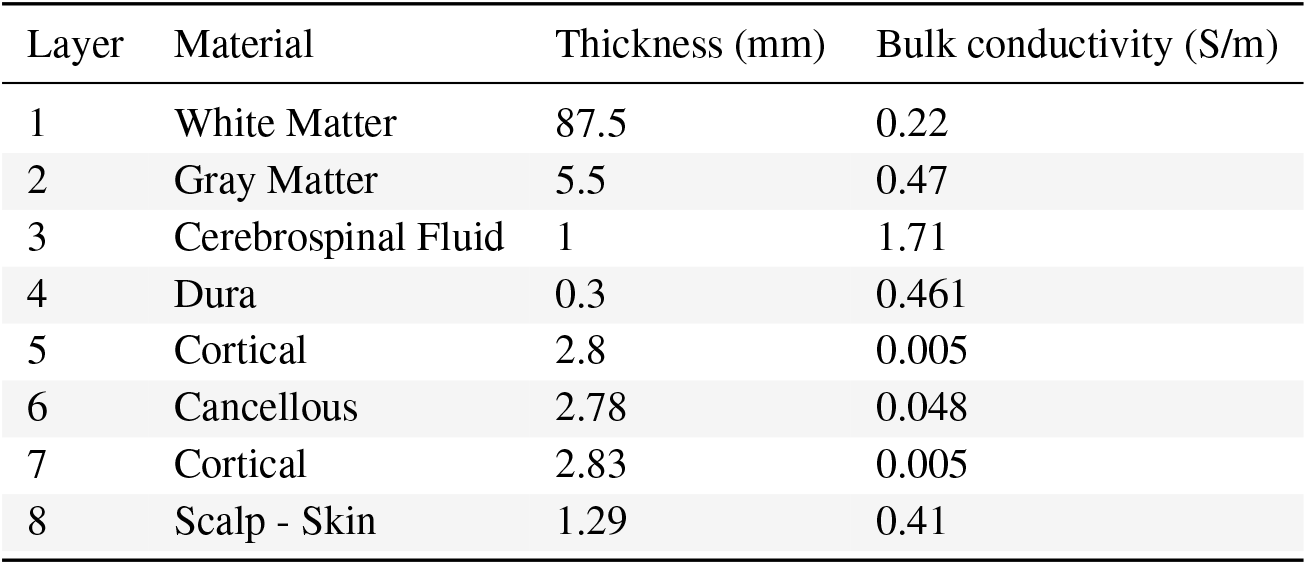
Electrical properties of the seven-layered spherical model.

| Layer | Material | Thickness (mm) | Bulk conductivity (S/m) |
| --- | --- | --- | --- |
| 1 | White Matter | 87.5 | 0.22 |
| 2 | Gray Matter | 5.5 | 0.47 |
| 3 | Cerebrospinal Fluid | 1 | 1.71 |
| 4 | Dura | 0.3 | 0.461 |
| 5 | Cortical | 2.8 | 0.005 |
| 6 | Cancellous | 2.78 | 0.048 |
| 7 | Cortical | 2.83 | 0.005 |
| 8 | Scalp - Skin | 1.29 | 0.41 |

The implant was modeled as a disk of insulating material with a bulk conductivity of zero, inserted between the top layer of the skull (cortical bone - layer 7) and scalp (layer 8) as shown in Fig. 2(a). Here one half of the spherical model of the head is presented, with the layer number corresponding to that of Table 2.1. The thickness of the insulating disk was set as 0.3 mm. The diameter of the disc was set as a variable with values of 4 cm, 5 cm, and 6 cm. These are shown in Fig. 2(b). The EEG signal source was modeled as a current dipole, with a magnitude of 0.1 mA and a dipole length of 1 mm. This is depicted in the schematic shown in Fig. 2(a) and FEM diagram in Fig. 2(d). The dipole was placed in the white matter region of the brain at variable locations relative to the implant ranging from 1 cm to 5 cm. The simulation was performed for the cases where the dipole is perpendicular to the implant plane and parallel to the implant plane.

**Figure 2:**
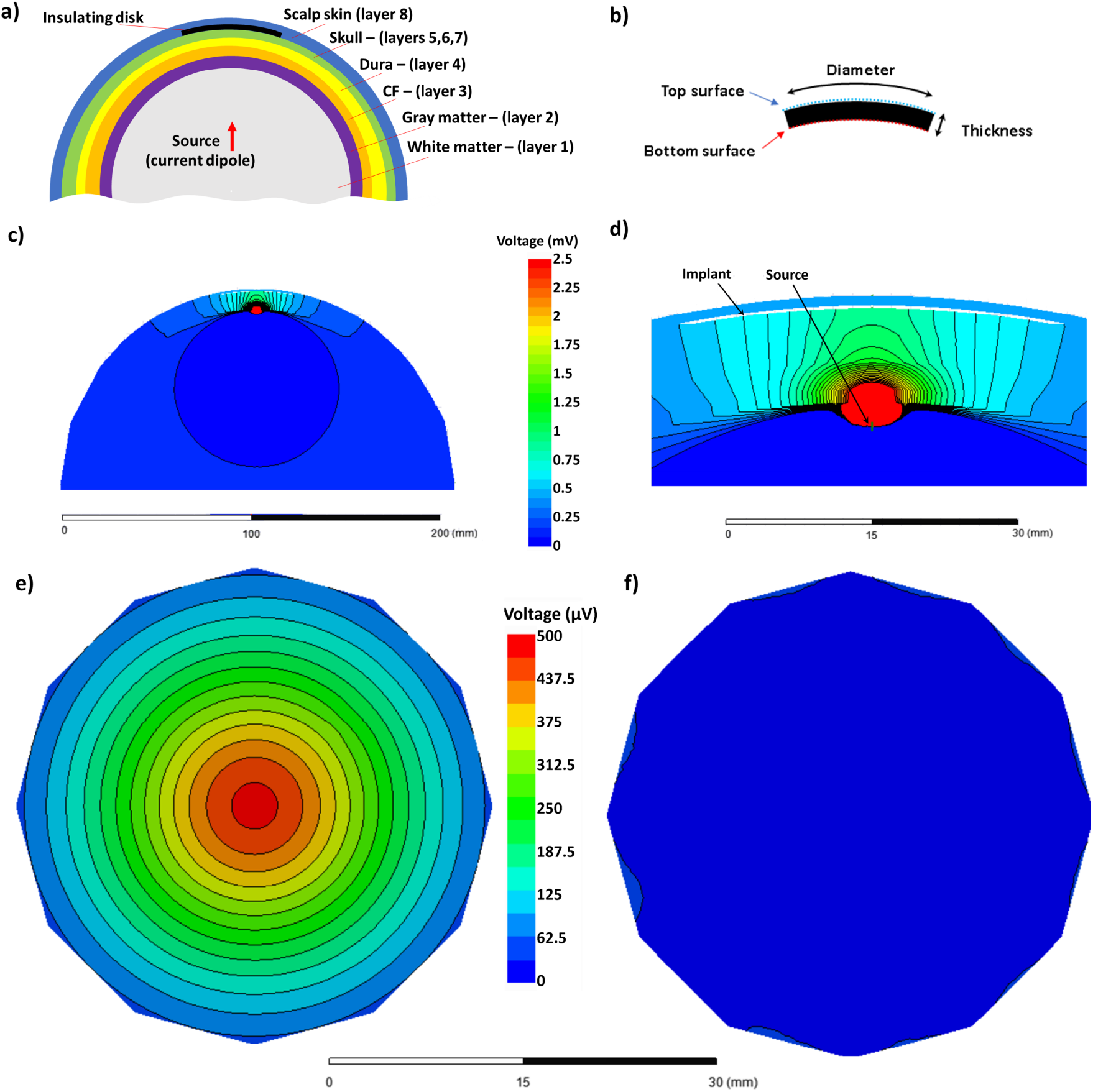
(a) Electrostatic finite element modeling setup showing the eight layers used in the model as well as the relative placement of the source (current dipole) and the insulating disk (implant model). (b) The insulating membrane’s cross-section is showing the top and bottom surfaces used to extract the differential voltage values. (c) A typical finite element method (FEM) simulation result highlighting the voltage variation. (d) Zoomed-in version of (c) highlighting the location of the implant and signal source. (e) and (f) voltage variation over the bottom and top surfaces of the insulating membrane. Throughout the FEM analyses, the voltage at the center of the insulating membrane, over a diameter of 0.1 mm, is used in the calculations.

The FEM mesh consisted of 464K tetrahedral elements from 20K nodes using a maximum element length of 0.5 mm at close proximity to the implant. The meshing and electrostatic simulation were performed using the Ansys Electronics Desktop 2019 R3.7 environment. The results we measured by analyzing the voltage difference between the top and bottom surfaces of the insulating membrane as shown in Fig. 2(b). Typical FEM outputs are shown in Fig. 2(c) to 2(f). One half of the spherical model of the head is shown in Fig. 2(c). The black lines show the equipotential lines. Fig. 2(d) is the zoomed-in version of Fig. 2(c) and highlights the location of the implant and the source. As mentioned, the diameter of the implant, source-implant separation and relative orientation was set as a variable in this study. Fig. 2(e) and (f) shows the voltage at the bottom and the top surfaces. of the implant, respectively. The difference between the top surface and bottom surface was used throughout this work.

### 2.2 Boundary element modeling

Though the finite element method (FEM) is helpful to study how our device is functioning, it is highly complex computationally and not ideal for experiments that we need to repeat several times with different configurations of dipole and electrodes. Besides FEM, we use the boundary element method (BEM) and head model with three surfaces: outer skin, outer skull, and inner skull surfaces using a small subset of the Fieldtrip Matlab toolbox developed by Klaus Linkenkaer-Hansen and German Gomez-Herrero. To better emulate the subgaleal electrodes’ performance, we placed them on the outer skull surface.

### 2.3 Phantom experiment

#### 2.3.1 Flexible PCB electrode design

To verify the performance of the newly proposed electrode configuration, a conventional flexible four-layer printed circuit board (PCB) was developed. The PCB consists of copper tracks (thickness of 18 *µ*m for inner layers and 26 *µ*m for outer layers) and polyamide substrate with a total thickness of 0.26 mm. Gold-electroplated (copper immersion gold, immersion thickness of 4 *µ*m) contacts were exposed on both sides of the PCB. The top recording electrode consisted of six circular pads with a diameter of 4 mm. These were organized in a circle with a diameter of 2 cm. The bottom recording electrode has an identical layout and placed in a concentric position relative to the top electrode. The polyamide substrate was extended beyond the PCB’s center to investigate the role of the insulating membrane’s diameter, with white lines denoting insulating membrane diameters of 3 cm, 4 cm, 5 cm, and 6 cm.

#### 2.3.2 Phantom specifications

We experimented with saline solution (0.9%) as a liquid phantom. Specifically, to make the saline solution, we dissolved 0.9 g of analytical-grade NaCl with 1,000 ml of distilled water. The saline solution was placed in a container depicted in Fig. 3(a). The container has a flat circular bottom with a diameter of 15.5 cm, a top rim with a diameter of 22.8 cm, and a height of 12.8 cm.

**Figure 3:**
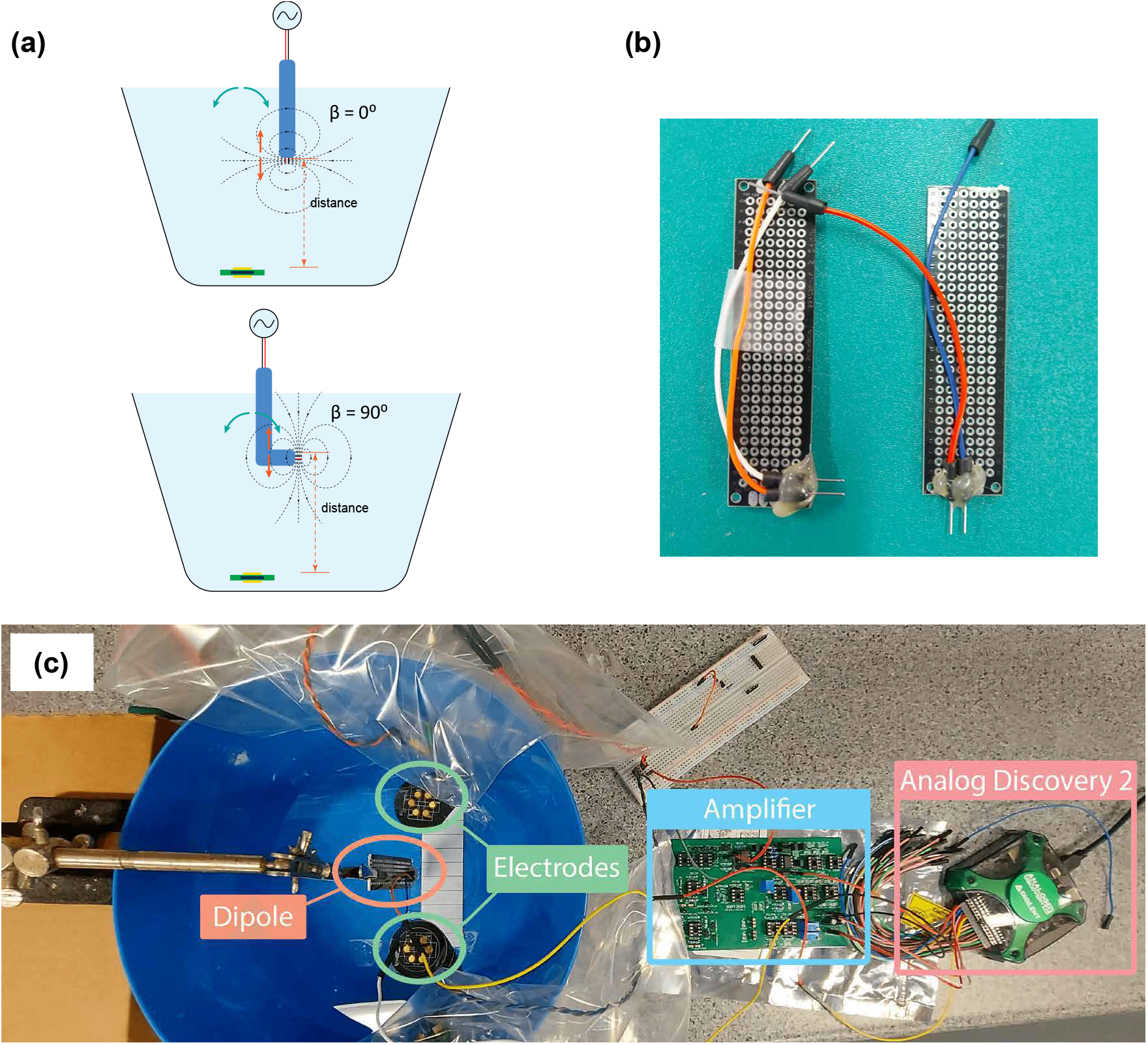
Experiment with a phantom. (a) Saline solution was used as a phantom model. The device with the top and bottom electrodes was placed at a 3 mm distance with the bottom of the tank. (b) The dipole electrodes were positioned 2.5 mm apart. The dipole configuration produces an electric field tangential or perpendicular to the surface of the electrodes. (c) It shows the setup and recording readout circuitry. Note that only a small part of the large amplifier board is used to host our low-noise amplifier components in this experiment

#### 2.3.3 Recording and signal source

The signal source was emulated using a voltage dipole consisting of two tinned copper electrodes with a separation of 2.5 mm. Two sets of dipole electrodes were used (see Fig. 3(b)). One of them generated a dipole perpendicular to the PCB plane, while the other generated a dipole in a parallel orientation relative to the surface of the flexible PCB. The electrodes were connected to a function generator (Analog Discovery 2), and a sinusoidal waveform with an amplitude of 100 mV and a frequency of 1 kHz was supplied.

The phantom setup is illustrated in Fig. 3(c). The flexible PCB was used to record the signal generated by the voltage dipole source. The flexible PCB was placed at the bottom of the tank with a gap of 3 mm to mimic the intended placement under the scalp. We measured the signal recorded with different distances and angles between the dipole and the device. As the signal recorded by the PCB electrodes is small, we employed a custom-made two-stage amplifier with a total gain of 80 dB. National Instruments Analog Discovery 2 board is used to supply signals to the dipole and readout the amplified signals.

### 2.4 Cadaver experiment

The phantom model provided an opportunity for a thorough assessment of the proposed solution from the geometric perspective. However, given that it only consists of saline solution, it lacked the complexity of a brain inside the head in terms of the various tissue types and their associated electrical properties. To address this, electrical measurements were obtained from the sensor implanted in a mouse cadaver. The Fig. 4 schematic summarizes the experimental setup used in the study. The measurements were undertaken using adult C57/Bl6 mice (experiments approved under RMIT Animal Ethics, AEC1933), aged approximately 10 weeks of age, immediately after they were killed using an overdose of pentothal (200 mg/kg). A dipole signal source was constructed as a needle electrode pair. A pair of enameled coated copper wires with a diameter of 0.3 mm were inserted in a 20-gauge needle (internal diameter 0.91 mm). The needle was backfield with epoxy resin (Skelleys Araldite 5 Minute Epoxy Adhesive). Abrasive sandpaper was used to polish the tip of the needle electrode pair to expose the electrode pairs. The needle was used to deliver a sinusoidal waveform (Amplitude 20 mV, Frequency range 1.3 Hz to 1 kHz) generated using a function generator (Keysight InfiniiVision DSO-X 3024A). The subcutaneous device consisted of two electrodes with the size of 2 mm*×*2 mm fabricated using 12 *µ*m thick brass sheets (GoodFellow). The electrodes were soldered to the enameled copper wires (300 *µ*m thick). They were attached at the center of a 12 *µ*m thick sheet of polyimide (GoodFellow) with a lateral dimension of 2 cm *×* 2 cm using the epoxy resin. The voltage difference between the two electrodes was measured using an oscilloscope (Keysight InfiniiVision).

**Figure 4:**
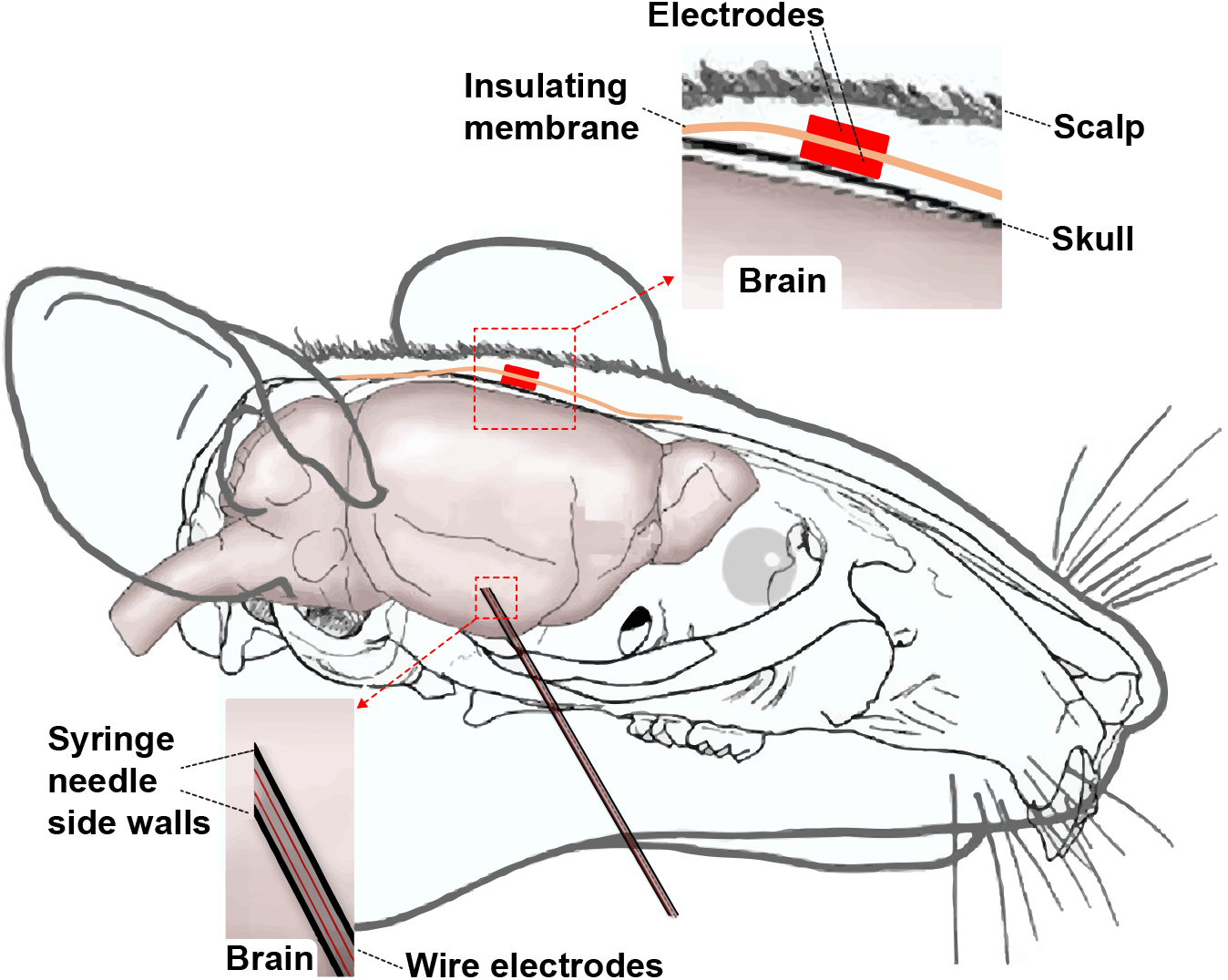
Electrical measurements with sensors implanted in a mouse cadaver. The subcutaneous implant consisted of two 2 mm *×*2 mm electrodes fabricated using 12 *µ*m thick brass sheets. They were attached at the center of a 12 *µ*m thick sheet of polyimide with a dimension of 2 cm *×* 2 cm using epoxy resin. The dipole signal source was constructed with a pair of enameled coated copper wires (diameter of 0.3 mm) inserted into a 20-gauge needle (internal diameter of 0.91 mm). The dipole signal had an amplitude of 20 mV and a frequency range of 1.3 Hz to 1 kHz.

The needle electrode pair (source) was inserted through the chin and up into the brain, approximately 1 cm. The subcutaneous electrode was inserted by making a 1 cm incision in the scalp and making a small pocket under the skin. Before the insertion, it became necessary to trim the polyimide (PI) sheet. The electrode was placed into the pocket, and the polyimide sheet smoothed down to contact the skull. The skin was closed to secure the electrode using four sutures. The minimum distance between the edge of the insulating PI and the electrode was 7 mm, and the maximum was 14 mm.

## 3 Results and Discussion

### 3.1 Top-bottom vs. side-by-side placement of electrodes

By putting electrodes on two sides of an insulator, we increase the effective distance between the two electrodes. In this section, we compare our top-bottom placement with the conventional side-by-side placement of electrodes under different configurations (see Fig. 5). To have a reasonable comparison, we put the two electrodes 6 cm center-to-center away from each other for the side-by-side placement. For the top-bottom placement, we used an insulator with a diameter of 6 cm, i.e., effective “center-to-center distance” would also be 6 cm.

**Figure 5:**
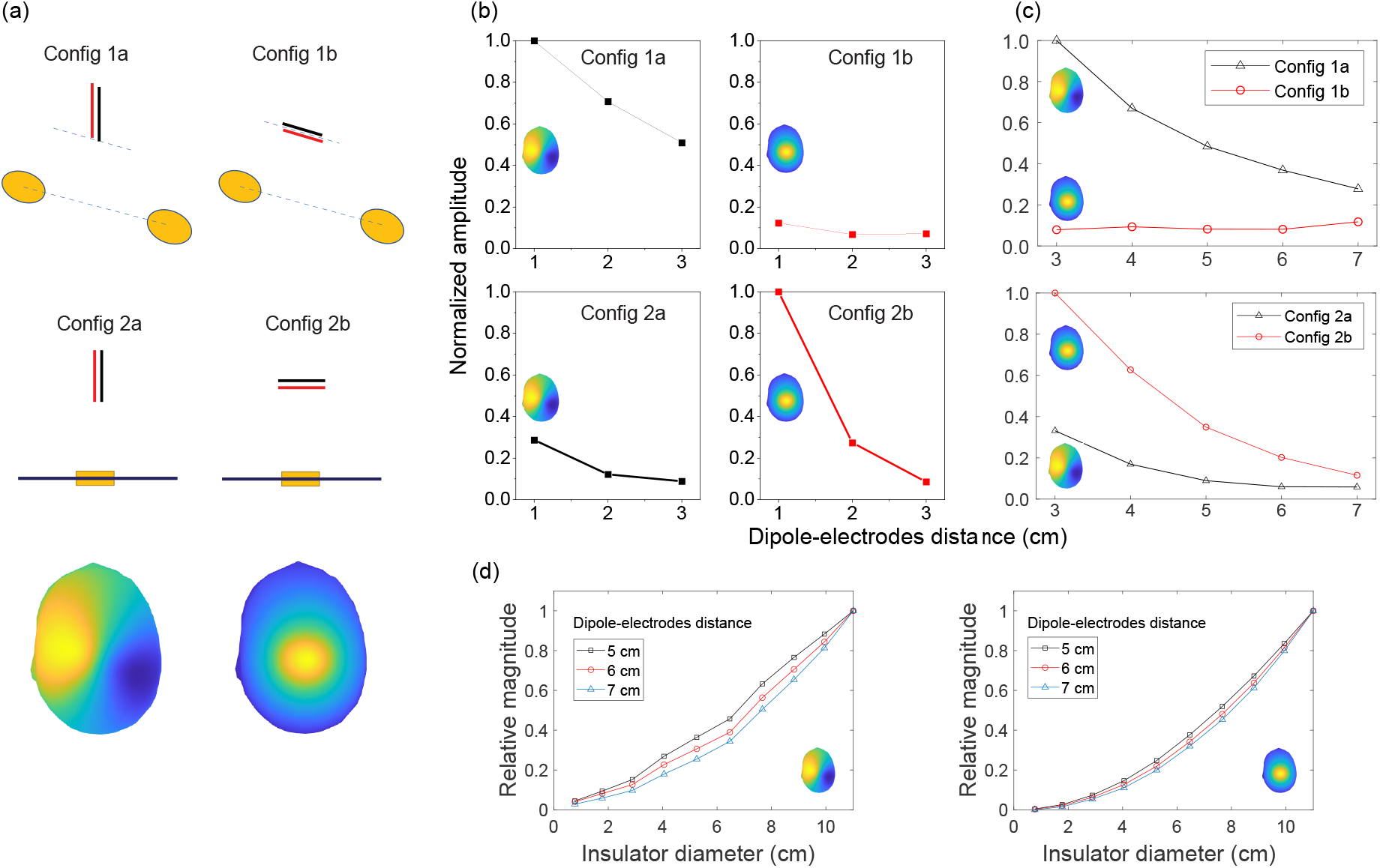
Comparison between side-by-side (configurations 1a and 1b) and top-bottom (configurations 2a and 2b) placements of electrodes in terms of recorded signals. (a) Dipole electrodes setups. Config 1a: tangential dipole is placed on top of the centerline of the electrodes. Config 1b: radial dipole is placed on top of the centerline of the electrodes. Config 2a: tangential dipole with top-bottom placement. Config 2b: radial dipole with top-bottom placement. (b) Measurements of recorded signals from the phantom experiment. Signals are normalized by dividing with the max amplitude per each electrode placement (side-by-side or top-bottom. (c) Matlab simulation results of recorded signals. Similarly, values are normalized by dividing with the max value per each placement. (d) Matlab simulation of the impact of the insulating membrane diameter to recorded signals when the dipole is tangential (left) and radial (right).

In electrophysiology, the dipoles create electric field components along three axes. This includes tangential components and radial components [38, 31]. While it is rare that the dipole in any orientation is placed right at the weakest point for signal sensing, but it is important to note shortcomings in our proposed structure and the conventional side-by-side electrodes. These configurations are presented in Fig. 5(a). As shown in Fig. 5(b), the amplitude of signals recorded using our electrode structure is a function of the distance between the source and electrodes, as is for the conventional side-by-side electrodes. While our electrode architecture is more sensitive to the radial dipole, the side-by-side electrodes are more sensitive to the tangential dipole, and it struggled to sense the signals, when the dipole is radial. In Fig. 5(c), we reported our Matlab simulation results for the corresponding phantom experiments. Note that in the head model, the distance between the brain, where dipole is located, and the electrodes on the outer skull surface is more than 2 cm. We performed the simulation with the dipole-electrodes distance starting from 3 cm onward. Fig. 5(d) demonstrates Matlab simulation results for our electrode architecture when we change the insulation layer diameter for two dipole-orientations and three dipole depths.

### 3.2 Impact of implant diameter

The role of insulating membrane diameter was assessed by comparing the signal amplitude recorded using the newly proposed electrode configuration against the signal amplitude recorded using conventional electrode configuration with the two electrodes. As expected, the reduction in the device insulating membrane diameter results in a decrease in the amplitude of the recorded signal. There is a solid and robust body of evidence for the distance dependency of surface EEG and experiments [39, 40]. The behavior that we observe in our device is, however, could be explained with the Laplacian EEG [41, 31].

#### 3.2.1 Laplacian EEG

Understanding conventional side-by-side EEG is very intuitive in terms of current flow between the two electrodes. The Laplacian EEG however involves a second-order partial derivative of a scalar potential distribution [41]. While a reliable estimation of the Laplacian could be challenging, its physical interpretation could help in understanding its link to our electrode architecture. Starting from the Coulomb’s Law and the interlink between the electric fields (**E**), generated by point charges, there will be an electric potential *V*_AB_ caused by of the field **E** between positions A and B. Continuing with the Gauss’s Law, the divergence of the electric field in a macroscopic sense (∇ · **E**), has components along *x, y* and *z* planes in Cartesian coordinates. A gradient relationship then relates **E** and *V*, **E** = −∇*V*. We can then see how the charge density relates to the potential by Poisson’s equation, which in a 2D approximation of the scalp can be written as

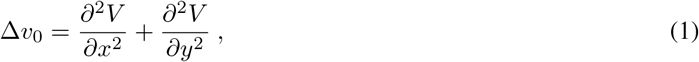

where Δ*v*_0_ is the Laplacian of *V* at the point 0, which is at the center of the finite difference five-point-method (FPM) calculation of Laplacian as described in [42] (see Fig. 6(a)). An approximation of the Laplacian can be achieved via

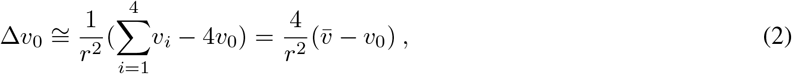

where 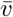 is the potentials on the ring or in it special condition on points *V*_1_ to *V*_4_ on the FPM [31, 42]. The distance between the center point, *v*_0_, and each of the other points (also the radius of the circle) is shown as *r*. The Eqn. 2 can be applied to the FPM model and also be extended to a bipolar configuration with a concentric disc and ring structure, as shown in Fig. 6(b). The estimation of Laplacian for the bipolar configuration, with the ring radius of *r*, is given in [42] as

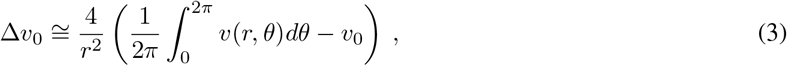

where 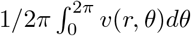 represents the potential on the ring 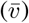 [42]. Taking the integral in Equ. 3 along the circle of radius *r*, shown in Fig. 6(b) and defining *X* = *r* sin(*θ*) and *Y* = *r* cos(*θ*), the Equ. 3 reduces to Equ. 2, and hence 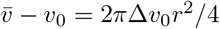. This illustrates an *r*^2^ relationship between the potentials. Fig. 6(c) shows the side cross-section of our proposed electrode architecture and we argue the combination of the top-electrode and the insulating membrane extension emulates the ring electrode in the bipolar configuration in Fig. 6(b). This is confirmed by a close agreement between the simulation of Equ. 2 and our simulation and experimental results, as shown in Figs. 5(b)-(d).

**Figure 6:**
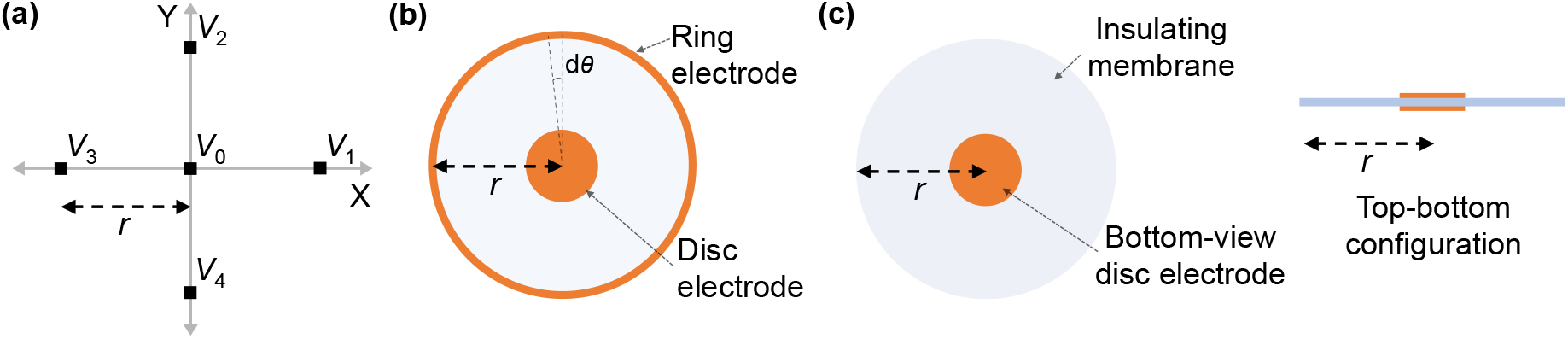
(a) Approximation of the Laplacian at the point *v*_0_ on a regular square grid plane using the five-point method (FPM). (b) A bipolar electrode configuration approximation of the FPM with a concentric disc and ring electrodes. (c) Bottom- and side-view of our proposed electrode architecture.

The bipolar structure is capable of estimating the Laplacian potential and is reported to be universally superior to the conventional side-by-side electrode system on SNR due to higher attenuation of common signals [43]. The Laplacian’s *µV/*cm^2^ property makes it substantially different from the conventional estimation of the electric field component tangent to the scalp surface, between two side-by-side electrodes. Usually, multiple electrode pairs in different directions are required to best sense 2D tangential components of the electric field. We discussed this further in the former part (3.1) and observed the shortcomings of our 2D systems and unreliability of the conventional 1D structures (see Fig. 1(a)) in sensing tangential and perpendicular components of the electric field.

### 3.3 Impact of source-implant separation

The separation between electrodes and signal source impacts the recording amplitude. With the increase in the source-electrode separation, the signal amplitude reduces and the coherence in recorded signals on different channels. This phenomenon has been widely reported for a variety of recording configurations and is a critical dipole feature that its potential drops as the inverse square of the distance in its surrounding medium [31, 41].

### 3.4 Impact of electrode diameter

The proposed implant configuration utilizes an insulating membrane to generate a difference between two electrodes placed at either side of the membrane. Fig. 7(a) depicts the voltage along the two surfaces of the implant. On the bottom surface, one of these surfaces is facing the dipole signal source, while the second surface, the top surface, is facing the scalp. The width of the insulating membrane with a diameter of 5 cm spans the length of *−*2.5 cm to 2.5 cm. Within this range, there is a contrast between the voltage bottom and top surfaces. The most considerable difference is observed at the center of the implant, marked as a length of 0 cm. As the diameter of the electrodes is increased, the voltage recorded by the electrodes would include locations that deviate from the optimum 0 cm length. These locations all exhibit smaller voltage recording at the bottom surface, as highlighted in Figure. 7(a). This means that an increase in the diameter of the recording electrodes would reduce the differential signal recording. This is particularly the case for the electrode facing away from the dipole source (i.e., the bottom electrode), where the variation between voltage and length is strongest.

**Figure 7:**
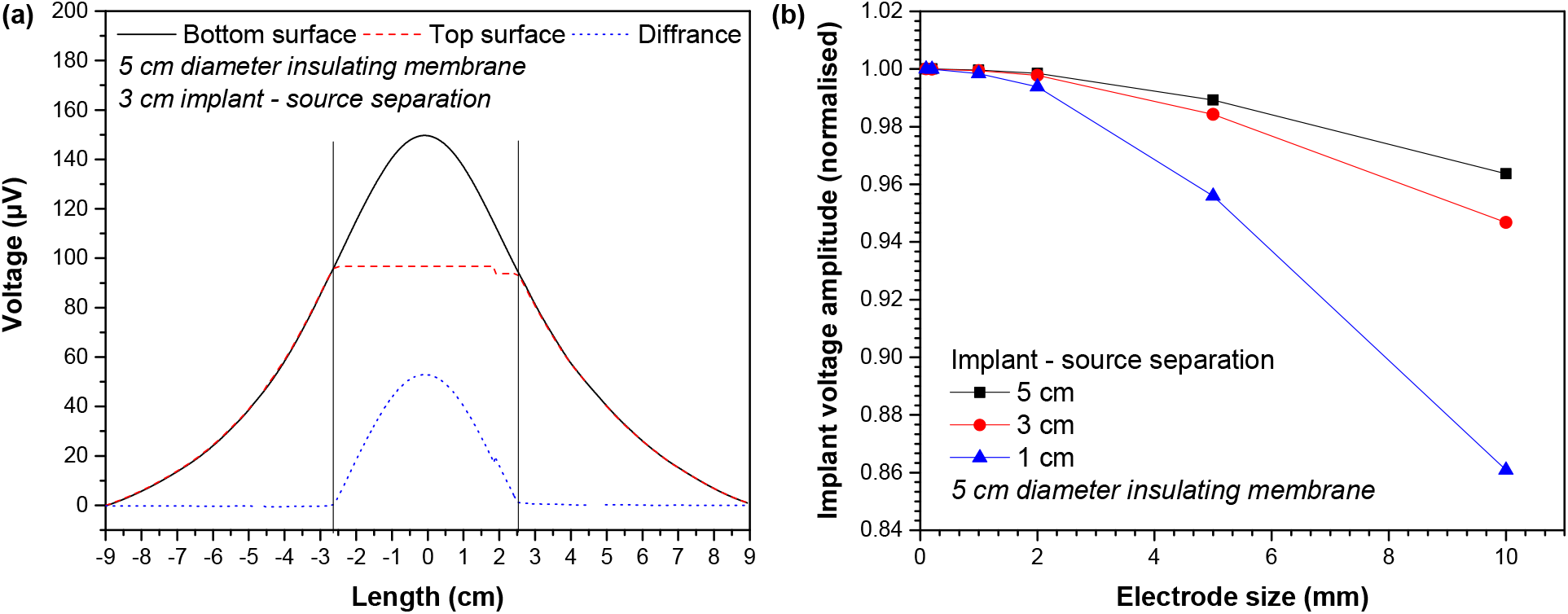
(a) Impact of electrode diameter. Voltage along two surfaces of the implant. Bottom surface is facing the dipole signal source while the top surface is facing the scalp. (b) Variation in the differential recording with different electrode sizes.

Fig. 7(b) highlights this by showing the variation in the differential recording (normalized) as a function of the electrode size. The results here are obtained from the FEM simulation, and the voltages are calculated based on the minimum voltage observed at the edge of the electrodes. As highlighted, an increase in the electrode size results in a reduction in the differential recording.

### 3.5 Impact of source ordination

The ordination of the source relative to the recording device would impact the voltage amplitude. The results presented in 3.2, 3.3, and 3.4 are based on the assumption that the source dipole is oriented perpendicularly relative to the implant’s surface. We appreciate that in many cases, this is may not be the case. As shown in Fig. 8, the normalized differential voltage recorded by the implant is larger in the case of perpendicular dipole orientation (zero degree) compared to parallel dipole orientation (90° degrees). While at the signal source separation of 1 cm, perpendicular dipole orientation yields a normalized differential recording of 1, whilst parallel dipole orientation produces a normalized differential recording of 0.4. However, as the implants or separation increases, the differential recordings in the cases of perpendicular and parallel dipole orientations deviate further. For example, active implants source separation of 5 cm, the normalized differential voltage recording in the case of for perpendicular dipole orientation is *≈* 2 *×* 10^*−*2^, whilst a normalized differential voltage recording of *≈* 10^*−*4^ is attainable in the case of parallel dipole orientation.

**Figure 8:**
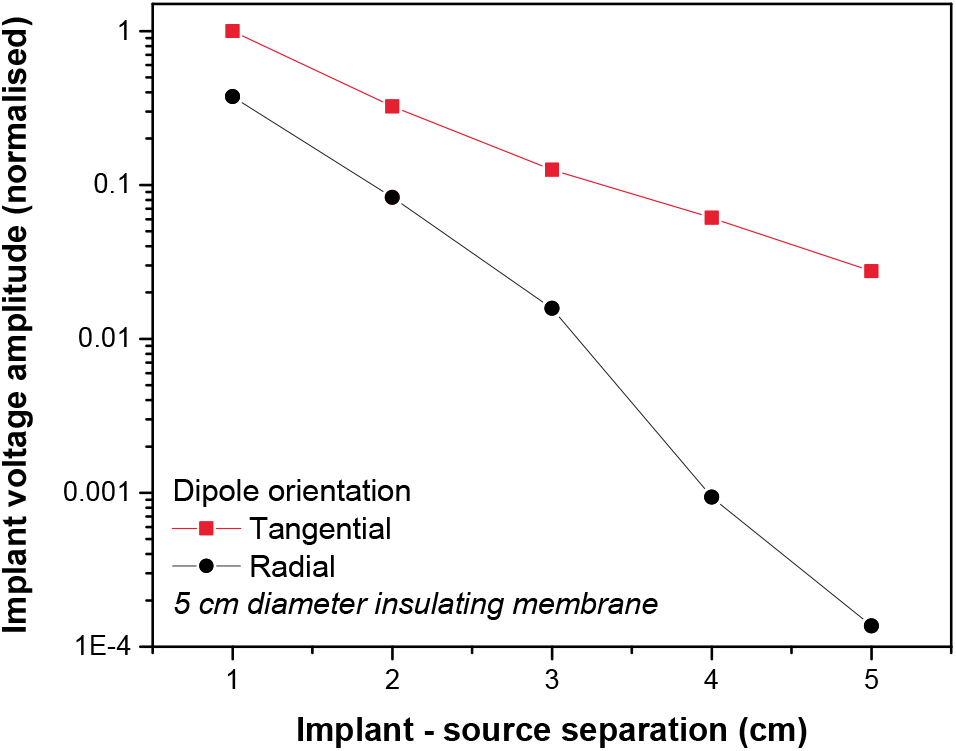
Impact of source ordination on signal recording. The normalized differential voltage recorded by the implant is larger in the case of radial dipole orientation compared to tangential orientation.

### 3.6 Cadaver experiment

Fig. 9 shows a summary of experimental results obtain from the animal cadaver experiment. The signal attenuation calculated as *V*_sense_*/V*_source_ is presented as a function of frequency. *V*_sense_ is the differential voltage recorded across the implanted recording device. *V*_source_ is the voltage across the dipole use as the signal source. As shown here, the implant is capable of high-quality differential recording across the physiologically relevant frequencies. Three orders of magnitude change in the frequencies, from 1 Hz to 1 kHz, results in one order of magnitude variation in the signal attenuation. This may be attributed to the tissue filtering effect, where differential biosignal recordings are generally biased towards higher frequencies because of the delay caused by the propagation of the signal across the tissue.

**Figure 9:**
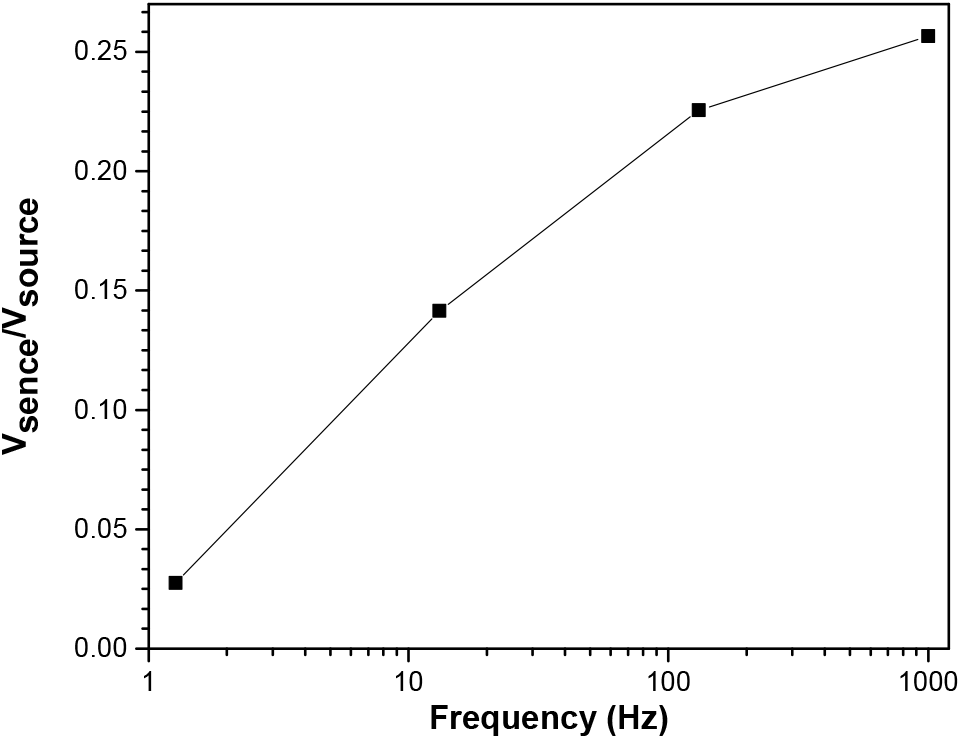
Recorded signal measurements across a frequency range of 1.3 Hz to 1 kHz from the mouse cadaver experiment.

## 4 Perspective and Future work

The ability to perform long-term EEG monitoring is currently very limited, and there are only a few potential viable solutions. Long-term EEG monitoring allows accurate measurement of seizure frequency and can complement short-term surface EEG in epilepsy diagnosis and management, and potentially implications for EEG recording protocols [44]. This study presents a novel solution with an electrode design that dispenses the need for a lead wire. We provided experimental and modeling evidence that a reliable readout with our novel electrode architecture is feasible. There are benefits, disadvantages, and remaining questions that we aim to raise in this section.

The golden standard of epilepsy diagnosis relies on surface EEG readings and its accurate and objective annotation [8]. Over the past two decades, there has been widespread use of EEG signals for seizure detection and seizure forecasting [10, 45]. Unfortunately, wearable EEG solutions are not designed for long-term use. They may cause skin injury and scarring, and infection. Highly invasive solutions such as intracranial EEG also may not be acceptable or justifiable for many participants [16, 46, 47, 48]. Subgaleal EEG (sgEEG) allows for an ultra-long-term EEG recording with high stability and data quality. There may be applications beyond epilepsy, such as brain-computer interfaces or gaming. It appears to be a safe and reliable technology when compared to other devices in development [7]. The technology could help personalize epilepsy treatments, track seizures in real-time and remotely, assist in titration of medication, or monitoring cerebral function in other conditions e.g., predicting hypoglycemia in diabetic patients and sleep disorders. Subscalp devices are reporting encouraging results in seizure tracking capability and are superior to self-reported patient seizure diaries.

### 4.1 Seizure counting/detection

The International League Against Epilepsy (ILAE) defines epilepsy as a brain disorder that generates (1) two unprovoked seizures that are more than 24 hrs apart, or (2) one unprovoked seizure with at least 60% risk of recurrence over the next ten years [2]. Accurate and objective seizure counting in the long term has important implications for the management of epilepsy [2]. Many decisions on the treatment of epilepsy or regulatory trials for approval of medications for epilepsy rely on seizure frequency diaries that are known to be inaccurate. In addition, the misdiagnosis or delayed diagnosis of epilepsy is still common and has serious consequences [49, 50]. False positives can lead to the inappropriate prescription of AEDs that result in adverse effects or worsening symptoms [50, 51]. This issue is compounded by societal inequities, as 80% of patients with epilepsy are amongst low to middle-income populations, and 75% of them do not receive any treatment [6]. The treatment gap can be attributed to inequities in distribution and access to services, stigma associated with the disease, lack of sufficient expert resources (neurologists), and an inadequate supply of modern AEDs.

One thing that makes the diagnosis of seizure difficult is that seizures are generally infrequent in adults. They may occur weekly, monthly, yearly, or with a gap of a few years. The EEG patterns during a seizure are also quite unique to each patient, and individualization of seizure detection and forecasting is necessary [52, 46]. Undetected or unreported seizures are common and, in turn, can affect the diagnosis, the assessment of the risk of recurrence and injury. It may also impact the risk of the individual performing functions such as driving. Following severe brain injury, post-traumatic epilepsy (PTE) may develop months or years following the insult [53]. Among many types of clinical epileptic seizures, non-convulsive seizures and non-convulsive status epilepticus are severe medical conditions that could lead to an emergency department and intensive care units (ICU) admission or death [54, 39, 40, 55]. Recording of non-convulsive seizures using non-EEG methods is challenging and remains a challenging task even for EEG-based solutions [56]. Time-limited and expensive hospital bedside physiological monitoring is the only tool available to clinicians at this time.

### 4.2 Seizure prediction and closed-loop neurostimulation

The Epilepsy Foundation’s 2016 Epilepsy Innovation Institute (EI^2^) community survey showed that the unpredictability of seizures is one of the most significant concerns for people living with epilepsy, their caregivers, and family [57, 52]. Circadian (days) and multidien (multi-days) seizure cycles have been studied with patient self-reported diaries and continuous intracranial EEG, which shows peaks in seizure cycles as long as 30 days apart [58, 16, 46, 59]. Whether the observed cycles are real and whether they can be linked to missed medication, mental and emotional states, the menstrual cycle, and the duration and severity of seizures need objective and ultra-long EEG data on a scale of months and years [60]. One observation is that mammalian physiology and behavior is widely influenced by light and other environmental factors, which means circadian or multidien studies require well-developed protocols [61].

Besides social stigma, discrimination, and a high prevalence of severe depression in people living with epilepsy, a recent productivity study in Australia indicated that just a 5% improvement in seizure control could result in more than AU$2 billion in economic benefit for the country [62]. One way of increasing seizure control or increasing the quality of life for patients living with epilepsy is to provide a reliable and personalized way to predict seizures or detect them early to intervene using neuromodulation for aborting seizures [63]. It may also reduce the use of medical resources and anxiety around seizures.

### 4.3 External interference, motion artifacts, and impedance imbalance

Ideal signal amplification of EEG requires rejection of any common interference potentials if they are equally coupled in the differential inputs. Extraneous electrical sources, such as power lines, introduce a common-mode noise over a wide range of AC line frequencies, 50/60 Hz. This interference can be significantly more significant than the EEG signal. This potential is a function of the area of loop enclosed by wires ^3^. The design of our device significantly reduces the area of this loop compared to conventional solutions (Fig. 1(a)).

Another source of interference is capacitive coupling between the AC line, and lead wires, and the body. The mechanism of this interference is by injecting displacement currents into the leads, which then could cause an imbalance depending on differences in electrode-tissue impedances. This impact could be mitigated by eliminating lead wires. Overall, the displacement current may not cause significant interference for the available solutions. The electrode-tissue impedance for subgaleal EEG should generally remain balanced with careful placement and positioning. In addition, existing solutions, which have a larger electrode-tissue surface, may reduce impedance but are susceptible to more displacement current.

Our approach in co-integration of electrodes and amplification within sub-mm physical proximity reduces the signal path from electrodes to our low-noise amplifier from a maximum of 20 cm in available cited examples (see Fig. 1(a)) to approximately 200 *µ*m while being fully balanced, this is a reduction by a factor of 1000 leading to a significant decrease in other lead wire related parasitic effects. The mains interference also reduces as the signal path between the electrode and amplifier input is smaller. This is the method is used in designing dry electrode surface EEG systems. Furthermore, our aim in using complementary metal-oxide-semiconductor (CMOS) application-specific integrated circuit (ASIC) will provide robust, energy-efficient, and superior noise performance compared to off-shelf components. We believe that the removal of leads could also significantly reduce transient wire motion artifacts. While subscalp wire motions are not expected to be as significant as surface EEG, micro-movements may still occur depending on the subject’s level of activity (e.g., exercise or sport).

One of the most important figure-of-merits in the front-end ultra-low-noise-amplifier’s custom design is the common-mode rejection ratio (CMRR). In the absence of compensation, CMRR can be limited by impedance imbalance and amplifier’s internal mismatch. In custom ASIC design, effective techniques such as continuous-time common-mode feedback circuit (CMFB), front-end AC-coupled chopper modulation amplifier, and digitally-assisted offset trimming can be used to deal with non-idealities while maintaining an ultra-low-power operation [64].

### 4.4 Physiological artifacts and implant placement

The Information content in EEG is rich and across a broader range of frequencies beyond 5-100 Hz. It is, however, highly contaminated with a range of physiological artifacts produced by the heart, eyes, ambulatory motion, and cranial and cervical muscles. It is known that electrical signals from skeletal muscles (e.g. temporalis) contaminate EEG from around 20 Hz with a significant presence at frequencies above 30 Hz (gamma range) [65]. Wider band EEG is essential to many applications, and therefore various techniques such as blind source separation, independent component analysis (ICA), wavelet analysis, canonical correlation analysis, beamforming, and surface Laplacian transforms may be used to reduce electromyography (EMG) contamination [66, 41]. The level of muscle signal contamination was known when paralysis of the superficial muscles was reported to reduce gamma range power by about 98% at some surface electrodes [66]. The EEG is susceptible to muscle artifacts even at rest [67]. The cranial and cervical muscles are affected by emotion, stress levels, and cognitive tasks [66].

Implant positioning described in the UNEEG SubQ’s surgical procedure user manual suggests that all positions will involve significant temporalis muscle artifacts [68]. The placement of the electrode in proximity to the temporalis muscle not only increases EMG artifact, but could also increases risk of damage to superficial temporal vessels and nerves. The feasibility of placing our proposed electrodes is subject to ongoing research and will be investigated with a large animal study.

For the surface EEG, scalp muscle artifact contamination at each electrode is different, and therefore, it can be assumed that the currently available subscalp EEG would face the same challenge. Studies show the relative EMG contribution of electrodes adjacent to the temporalis muscles may be as high as 200 times the EEG, as compared to central electrodes where EMG artifact can be a factor of 10 to that of EEG [66, 67]. Major electromyographic artifacts are also reported in subscalp EEG, such as UNEEG’s SubQ device, with a different level of EMG contamination in each of their two electrode contacts due to differences in proximity to the temporalis muscle [29].

Expanding on the discussion in Section 3.2, the conventional surface electrodes need to be at least *∼*20 mm apart for sensing EEG, but the volume conduction through tissues of the head results in a reduction of the effective spatial resolution [40]. During fast repetitive movement-related EEG potentials, adjacent disc electrodes have highly correlated signals [42]. A change in the design of these electrodes to a tri-polar concentric ring electrode (tCRE) results in a significant reduction in the common potentials induced by movement [43]. The tCRE is reported to have a unique ability to enhance EEG signal quality, spatial resolution, SNR, source localization, and reducing EMG contamination. These electrodes can then be successfully used in seizure detection and to record high-frequencies in EEG [69, 70, 71, 72]. A tCRE is directly measuring the Laplacian potential, which is equivalent to the second spatial derivative (surface Laplacian) of the scalp electrical signals [41]. This is then followed by assigning different weights to each of the electrode rings in a linear equation that can be computed by the hardware [43].

The surface Laplacian transform (SL), with the unit of *µ*V/cm^2^, estimates the local current flux through the skull into the scalp and is widely reported to achieve an improved spatial selectivity and SNR over conventional surface potential measurement (unit of *µ*V) [73]. At the algorithmic level, a mix of ICA and SL is shown to significantly reduce EMG artifacts of the EEG in all frequencies [74]. While there are debates around the theoretical justification of surface Laplacian, it remains a concise and simple representation of the field topography and a physiologically meaningful transformation [31].

In our device, the combination of the top electrode and the extended insulating membrane emulate the outer ring of a bipolar common-centric design, with the bottom-electrode being its center disc. This is an emulation of the bipolar Laplacian structure [43]. The presented emulation is aligned with the theory behind the finite difference five-point-method (FPM) [31] (see Fig. 6(a)).

Dipoles in the cortex are usually perpendicular to the surface [31]. Like typical surface potential sensing, surface Laplacian is more sensitive to radial dipoles and provides a unique feature of much higher sensitivity than the surface potential to the superficial cortical sources, which could be the case in the case of focal epilepsy for instance. Such feature provides a further advantage, when the objective is source localization. The surface Laplacian spatial resolution to source signal is substantially higher, relative to the scalp potentials, which means smaller dimensions for sensing can be used [31].

### 4.5 Clinical trials of subscalp EEG

Major clinical trials of subscalp devices include trial registrations NCT04526418, NCT04513743, NCT04061707, NCT03465189, and ACTRN12619001587190 [75, 76, 77, 78, 79]. We are particularly interested in NCT03465189, in which a subcutaneous Medtronic’s heart monitoring device, Reveal LINQ loop-recorder, was re-purposed for brain monitoring. In this trial, the device is used in epileptic patients, and while results are not reported in detail, it is reported to be compared with the gold standard clinical recordings. This feasibility study is reported successful for epilepsy diagnosis, monitoring, and management. This battery-powered loop recorder and similar devices have loop memories that continuously record and overwrite data. It is unclear whether the LINQ device was placed subcutaneously or it was paired with lead wires placed subcutaneously. It is also unclear whether the device’s electronic part was modified for this study.

### 4.6 Implant thickness

Implant encapsulation technologies, as well as the necessary circuits for high-performing electronics, stimulation, or recording, have sufficiently demonstrated their capabilities for realizing ultra-thin implants that sit on the human retina [80, 81]. The entire system can be encapsulated in a rigid body with a size of a US cent coin, with a thickness of around 1 mm, making its rigid part *∼*300 *µ*l of volume. The flexible insulating membrane extension around the rigid part makes the total implant volume to *∼*1,000 *µ*l. The packaging is transparent (or could be made transparent) to near-infrared light and/or electromagnetic waves, making it possible to have an RF-link for power delivery and data telemetry or an optical link for power delivery and RF data telemetry.

### 4.7 Conclusion

Subgaleal placement of EEG electrodes has its advantages and disadvantages. It requires surgical implantation, has a lower spatial resolution in terms of the number of electrodes, and requires prior knowledge of optimal position for the electrode placement (e.g., the best location for recording seizure activity). However, it is the only known solution that allows minimally-invasive ultra-long-term EEG that meets the clinical needs such as epilepsy diagnosis. It will also allow researchers to take on the grand challenge of seizure prediction. Subgaleal EEG promises higher quality signal than surface EEG and is more stable. It is much less invasive and safer than intracranial electrodes. An electrode design would be preferable if it allowed: Placement without general anesthesia (using local anesthesia). A smaller implant for better levels of comfort and wound healing. The absence of leads that may be prone to breaking or failing. In this work, we have introduced a novel architecture of electrodes to be placed in the subgaleal space. The small dimensions of the proposed electrodes are also ideal given the spherical shape of the skull and the thin, soft tissue of the scalp. We have demonstrated that the back-to-back placement of electrodes allows a marked reduction in the size of the rigid part of the implant. We believe that this is the first time that it has been shown that the sensing distance could be decoupled from the physical distance between electrodes. Further research will be needed to address whether the electrode-tissue impedance imbalance can be addressed with proper readout circuitry, and the implant can effectively reject EMG artifacts.

## Data Availability

Appropriate detail of analysis and tools as well as methods in experiments, simulation, and modeling are provided in the paper.

## 5 Contributions

AA and NDT contributed equally to this study. OK, AA and AN developed the idea and the concept. OK led the research and development. AA, NDT, BF, and OK performed the experiments. AA, NDT, and OK constructed instrumentation. AA and OK performed, discussed and analyzed modeling. AA and BF constructed and conducted cadaver experiment. All authors contributed to writing the manuscript.

## 6 Ethics declarations

Animal experiments is performed under approved RMIT Animal Ethics AEC1933.

## 8 Acknowledgement

OK acknowledges the support provided by The University of Sydney through a SOAR Fellowship. Authors acknowledge insights from Dr Mohammad Mirkhalaf, from the ARC Training Centre for Innovative BioEngineering, about bone properties and relevant experiments.

## Footnotes

3 The law of magnetic induction.

